# Quantitative Radiomic Features from Computed Tomography Can Predict Pancreatic Cancer up to 36 Months Before Diagnosis

**DOI:** 10.1101/2022.02.18.22271190

**Authors:** Wansu Chen, Yichen Zhou, Vahid Asadpour, Rex A Parker, Eva Lustigova, Eric J. Puttock, Bechien U Wu

**Author notes:** **Correspondence:** Wansu Chen, Ph.D., Department of Research and Evaluation, Kaiser Permanente Southern California, 100 S Los Robles, 2nd Floor, Pasadena, CA 91101. **Address where the work was conducted:** 100 S Los Robles, 2nd Floor, Pasadena, CA 91101. **Guarantor of the article:** Dr. Wansu Chen is accepting full responsibility for the conduct of the study. She had access to the data and have control of the decision to publish. **Specific author contributions: Wansu Chen**: Conceptualization, Methodology, Software, Validation, Investigation, Resources, Writing – original draft, Writing – review & editing, Visualization, Supervision. **Yichen Zhou:** Methodology, Software, Validation, Formal analysis, Investigation, Data curation, Writing – review & editing, Visualization. **Vahid Asadpour:** Methodology, Software, Formal analysis, Investigation, Data curation, Writing – review & editing. **Rex A Parker:** Conceptualization, Methodology, Validation, Investigation, Data curation, Writing – review & editing. **Eva Lustigova:** Conceptualization, Validation, Investigation, Writing – review & editing, Supervision. **Eric Puttock:** Validation, Investigation, Writing – review & editing. **Bechien U Wu:** Conceptualization, Methodology, Validation, Resources, Writing – review & editing, Supervision, Funding acquisition.

## Abstract

**Objectives:** Pancreatic cancer (PC) is the 3^rd^ leading cause of cancer deaths. We aimed to detect early changes on computed tomography (CT) images associated with pancreatic ductal adenocarcinoma (PDAC) based on quantitative imaging features (QIF).

**Methods:** Adults 18+ years of age diagnosed with PDAC in 2008-2018 were identified. Their CT scans 3 months-3 years prior to the diagnosis date were matched to up to two scans of controls. Pancreas was automatically segmented using a previously developed algorithm. 111 QIF were extracted. The dataset was randomly split for training/validation. Neighborhood and principal component analyses were applied to select the most important features. Conditional support vector machine was used to develop prediction algorithms. The computer labels were compared with manually reviewed CT images 2-3 years prior to the index date in 19 cases and 19 controls.

**Results:** 227 scans from cases (stages: 35% I-II, 44% III-IV, 21% unknown) and 554 matched scans of healthy controls were included (average age 71 years; 51% females). In the validation dataset, accuracy measures were 94%-95%, and area under the curve (AUC) measures were 0.98-0.99. Sensitivity, specificity, positive predictive value, and negative predictive values were in the ranges of 88-91%, 96-98%, 91-95%, and 94-96%. QIF on CT examinations within 2-3 years prior to index date also had very high predictive accuracy (accuracy 95-98%; AUC 0.99-1.00). The QIF-based algorithm outperformed manual re-review of images for determination of PDAC-risk.

**Conclusions:** QIF can accurately predict PDAC on CT imaging and represent promising biomarkers for early detection of pancreatic cancer.

**WHAT IS KNOWN:**

- Pancreatic cancer is the 3^rd^ leading cause of cancer deaths.
- Early detection of pancreatic ductal adenocarcinoma (PDAC) is difficult owing to lack of specific symptoms or established screening.

**WHAT IS NEW HERE:**

- Quantitative imaging features (QIF) of pre-diagnostic CT scans can accurately predict PDAC in 3-36 months prior to diagnosis (accuracy 94-95% and AUC 0.98-0.99).
- QIF on CT examinations within 2-3 years prior to cancer diagnosis also had very high predictive accuracy (accuracy 95-98%; AUC 0.99-1.00).
- The QIF-based algorithm outperformed manual re-review of images for determination of PDAC risk.

## INTRODUCTION

Pancreatic cancer is the third leading cause of cancer deaths in the United States with 48,220 estimated deaths in 2021.^1^ The 5-year survival in 2012-2017 was only 10.8%.^1^ Pancreatic ductal adenocarcinoma (PDAC) is the most common form of pancreatic cancer accounting for up to 90% of all cases. Early detection of PDAC is difficult owing to lack of specific symptoms or established screening. As a result, nearly 50% of cases have distant metastases at the time of diagnosis. Given the challenges with early detection in PDAC, the Scientific Framework for PDAC issued by National Cancer Institute called for an evaluation of longitudinal screening protocols including imaging biomarkers for patients at high risk for PDAC.^2^

A key step for early detection is the ability to identify precursors of PDAC on conventional cross-sectional imaging. Abnormalities of the pancreas such as main duct dilatation may be early indicators of PDAC and can be detected on computerized tomography (CT) with a high degree of reproducibility.^3-5^ However, these findings are often non-specific and improved methods are needed to identify accurate and reliable early indicators of pancreatic cancer on cross-sectional imaging.

Automated radiomic analysis of quantitative imaging features^6^ (QIF) abstracted directly from cross-sectional imaging offers a promising opportunity to objectively identify precursor findings related to pancreatic cancer. Radiomic analysis has been used to predict survival of PDAC patients^7-15^, differentiate functional abdominal pain, recurrent acute pancreatitis, and chronic pancreatitis^16^, and distinguish autoimmune pancreatitis from PDAC.^17^ One study attempted to classify PDAC cases from normal pancreas based on QIF of CT images after cancer diagnosis;^18^ however, the ability to identify precursor lesions prior to cancer development remains a key step to early detection. A review including 70 studies concluded that “ radiomics of the pancreas holds promise as a quantitative imaging biomarker of both focal pancreatic lesions and diffuse changes of the pancreas”.^19^ However, studies utilizing QIF to predict PDAC are limited.

The objective of the present study was to determine the ability of radiomics-based direct image analysis to identify changes in the pancreas on cross-sectional imaging associated with the subsequent development of pancreatic cancer. Specifically, we sought to develop automated computer algorithms to identify important QIF and subsequently assess performance for prediction of pancreatic PDAC overall and within time-specific intervals prior to development of cancer.

## METHODS

### Study design and setting

We conducted a nested case-control study, including cross-sectional abdominal CT images from health plan enrollees of Kaiser Permanente Southern California (KPSC). KPSC is an integrated healthcare system that provides comprehensive healthcare services for more than 4.8 million enrollees across 15 medical centers and 250+ medical offices throughout the Southern California region. KPSC health plan enrollees are broadly representative of the Southern California population at-large in terms of diversity with respect to race/ethnicity, socioeconomic status and other demographics.^20^ The study protocol was approved by the KPSC’s Institutional Review Board.

### Eligible study subjects and CT scans

We identified adults 18+ years of age diagnosed with PDAC in 2008-2018 (index date, t_0_) based on the KPSC Cancer Registry by using the Tenth Revision of International Classification of Diseases, Clinical Modification (ICD-10-CM) code C25.x and histology codes listed in Supplemental Document 1. The KPSC Cancer Registry is a prospectively maintained registry and part of the Surveillance, Epidemiology, and End Results (SEER) reporting program. Patients with history of acute or chronic pancreatitis, history of pancreatectomy or did not have at least 12 months of health plan enrollment prior to t_0_ were excluded (a gap of ≤45 days was allowed). CT scans 3 months-3 years prior to t_0_ of the eligible PDAC cases were obtained and were matched to up to two scans of controls by age, gender, race/ethnicity, CT contrast status and year of scan (±2 years). Controls met the same eligibility criteria above but were pancreatic cancer free up to t_0_ of the matched cases. All the CT scans were manually reviewed to remove those that did not capture the entire pancreas or had formatting errors. The resolution of scans was 512 × 512 pixels with slice thickness in the range of 2.5-5 mm.

### Characteristics of study subjects

Patient demographics, behavioral, and clinical characteristics on t_0_ or in the 12 months prior to t_0_ were extracted. Scan indications and the associated diagnosis for the visit were also captured. The tumor size was determined by manual review of radiology notes within the window of 3 months at the time of cancer diagnosis. The definitions of the clinical features are described in Supplemental Document 2.

### Pancreas segmentation

The method to automatically extract the volumetric shape of the pancreas was previously described.^21^ When the same method was applied to the images of the current study, we started from the estimated parameters previously derived and adjusted them to fit the data of the current study (see the method of enhancement in Supplemental Document 3). To evaluate the performance of the adjusted algorithm, we calculated Dice Similarity Coefficient (DSC) based on 9 randomly selected scans of PDAC cases by comparing the automated segmentation and that of manually delineated by the study radiologist (RP).

### Extraction of QIF

First, we normalized center and width of intensity window of all the scans. 111 previously validated QIF^22^ (Supplemental Document 4) were extracted from the segmented areas of pancreas using MATLAB software.^23^ Finally, the QIF were standardized such that they all had zero-mean and one standard deviation.^24^

### Algorithm development and validation

#### Preparation of training and validation datasets

The entire dataset was randomly split for training (50%, DS1) and validation (50%, DS2). DS2 was further divided into 5 subsets based on the temporal distance between the scan date and t_0_ (or t_0_ of the matched case): 3-6 months, 6 -12 months, 12-18 months, 18-24 months, and 24-36 months. By design, there is no overlap between training and validation datasets, and an individual scan was included in only one dataset.

#### QIF selection method

Two competing methods of feature selection were applied and compared based on DS1. The neighborhood component analysis (NCA) algorithm is a non-parametric method aiming for maximum prediction accuracy^25,26^. Using a regularization parameter (*λ*), the process of NCA learns feature weights by minimizing the expected leave-one-out classification accuracy. To avoid over-fitting, we tuned the regulation parameter *λ* based on 5-fold cross validation. The *λ* corresponding to the minimum classification loss was selected as the best. QIF with a weight greater than 2% of the maximum weight under the best *λ* was deemed important^26^ In addition, we applied a second approach to feature collection: principal component analysis (PCA) to transform all 111 QIF into linear combinations that are orthogonal to one another.^27^ Principal components with eigenvalues ≥1 were considered important. The analyses were performed by function fscnca in MATLAB^28^ and prcomp function in R^29^ respectively.

#### Algorithm development

Conditional support vector machine (SVM)^30^ was applied to develop prediction algorithms based on DS1. The reason for using “ conditional” SVM is to account for the paired structure of the matched data. More specifically, we centered the within pair data by its mean.^30^ For example, for a specific feature, the values were 0.4 and 0.6, respectively, from the images of a case and one of his/her controls, the centered values became -0.1 and 0.1. SVM is a high-performing non-linear classifier to map input data into higher dimensional space with the purpose of better ability of data separation^31^ Moreover, SVM can ignore outliers. The kernels functions used in the study included Gaussian, linear, and sigmoid.^32,33^ The two hyperparameters were tuned based on 5-fold cross-validation (Supplemental Document 5).

#### Algorithm validation

The performance of the prediction algorithms was evaluated using sensitivity, specificity, positive predictive value (PPV), negative predictive value (NPV), accuracy and area under curve (AUC) based on DS2. Accuracy was defined as the total number of correctly predicted individuals divided by the total number of patients. The validation was performed using the entire validation dataset and repeated in each of the 5 validation subsets.

#### Exploratory analyses

Because the selection of QIF by NCA may vary largely from one dataset to another, we conducted exploratory analyses to understand how the instability of QIF selection may impact the performance of final prediction algorithms (Supplemental Document 6).

### Clinical evaluation

A blind manual re-review of scans of 100% cases and 50% controls in the validation dataset 24-36 months prior to t_0_ was performed by the study radiologist (RP) to determine the risk of PDAC. Patients were classified as **low risk** if they only had diffuse atrophy, smaller cyst, simple cyst, loss of normal lobulation, or mild diffuse duct dilatation/prominence, or did not have any obvious morphological features. Patients with complex cyst, cyst larger than 2 cm, loss of normal lobulation of contour, or having two or more low risk features were considered having **medium risk**. Those with solid mass, focal abnormal enhancement, focal duct stricture, or focal/segmental atrophy were deemed **high risk**. The reviewer was not informed about the computer labels or case/control status at the time of review. The manual risk assessment was compared with the assigned computer labels (high risk “ 50%+” vs. low risk “ <50%”) based on the prediction algorithm developed with Gaussian kernel function and PCA.

### Statistical analysis

Characteristics of cases and controls were compared by using a conditional logistic regression model to account for the matched design. All the analyses were performed using SAS^34^ except for the MATLAB or R packages^35^ mentioned previously.

## RESULTS

### Characteristics of the Study Cohort

The study included 277 scans of PDAC cases and 554 matching scans from controls (Figure 1). 35% cases were stages I-II, 7.6% stage III, 36.8% stage IV, and 21% stage unknown at the time of diagnosis (Table 1). The median tumor size was 3.3cm (IQR 2.4-4.2) in the 148 cases with known information. On the average, the patients were 70.8 years of age and 50.5% were women. 47.3% were non-Hispanic whites, 26.7% were Hispanic, 18.4% were African American, and 7.6% were Asian/Pacific Islanders (Table 1). Ever tobacco use was frequent in both cases (57.1%) and in controls (51.7%) (p=0.14). Family history of pancreatic cancer, and diabetes were more common in cases than in controls (Table 1). Body mass index and weight change in one year were comparable between cases and controls. Among specified scan indications and associated diagnosis for the visit, abdominal pain, other pain, and GI problems appeared most frequent (Supplemental Document 7).

**Table 1.**
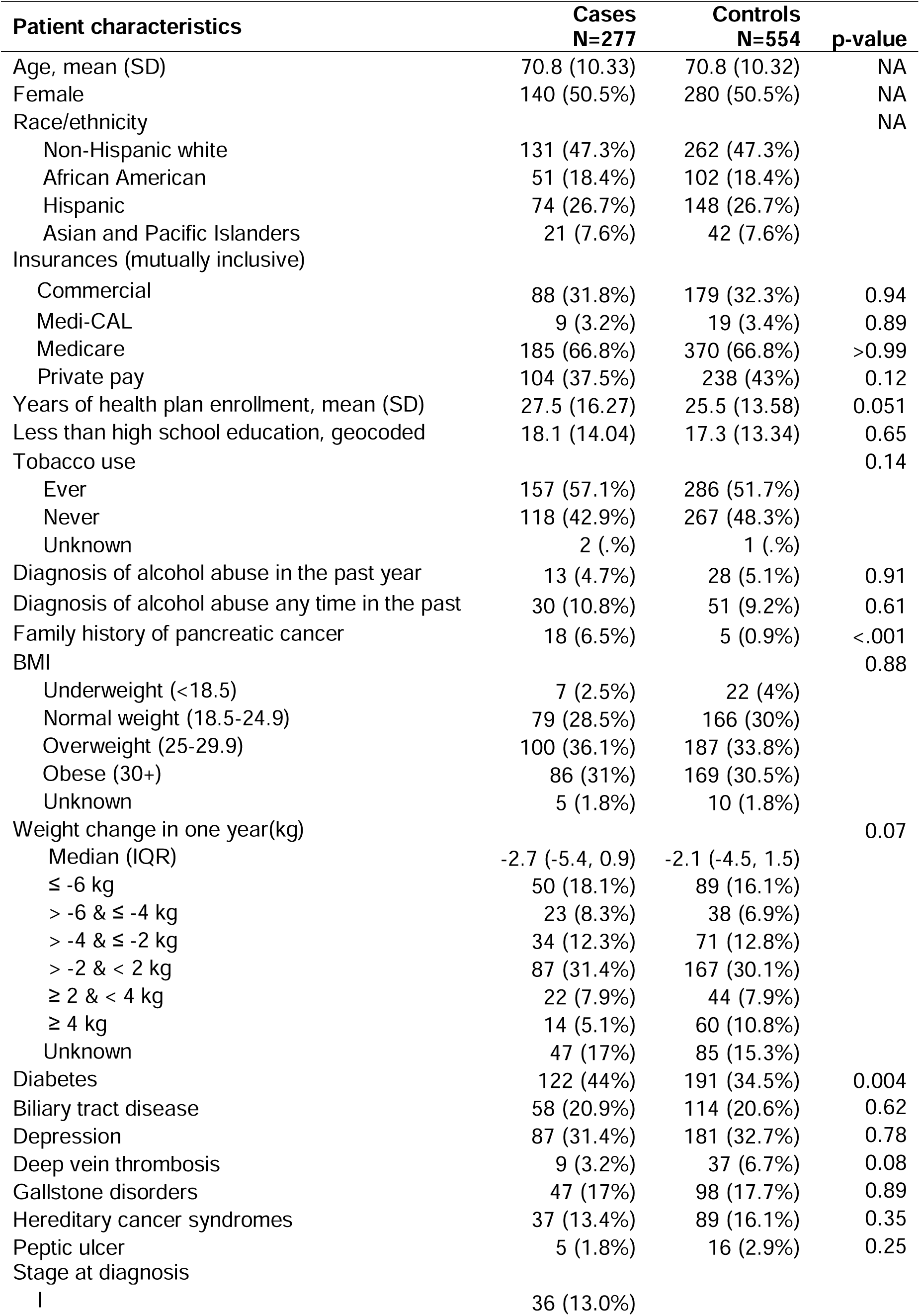
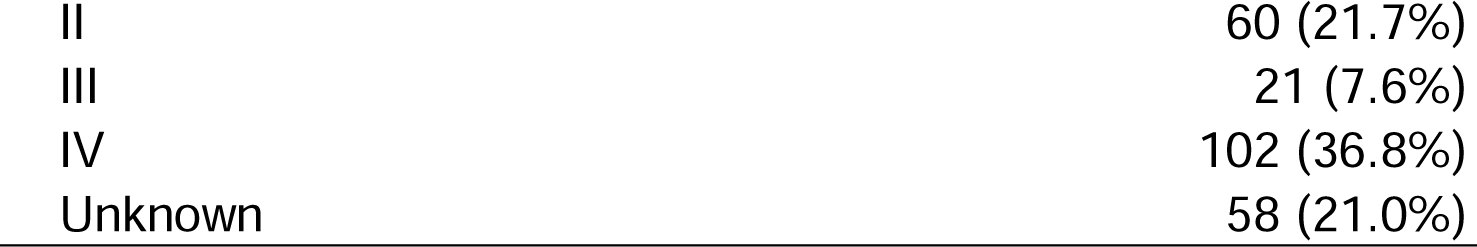
Characteristics of study subjects at baseline by case and control status, n (%) unless otherwise stated.

| Patient characteristics | Cases<br>N=277 | Controls<br>N=554 | p-value |
| --- | --- | --- | --- |
| Age, mean (SD) | 70.8 (10.33) | 70.8 (10.32) | NA |
| Female | 140 (50.5%) | 280 (50.5%) | NA |
| Race/ethnicity |  |  | NA |
| Non-Hispanic white | 131 (47.3%) | 262 (47.3%) |  |
| African American | 51 (18.4%) | 102 (18.4%) |  |
| Hispanic | 74 (26.7%) | 148 (26.7%) |  |
| Asian and Pacific Islanders | 21 (7.6%) | 42 (7.6%) |  |
| Insurances (mutually inclusive) |  |  |  |
| Commercial | 88 (31.8%) | 179 (32.3%) | 0.94 |
| Medi-CAL | 9 (3.2%) | 19 (3.4%) | 0.89 |
| Medicare | 185 (66.8%) | 370 (66.8%) | >0.99 |
| Private pay | 104 (37.5%) | 238 (43%) | 0.12 |
| Years of health plan enrollment, mean (SD) | 27.5 (16.27) | 25.5 (13.58) | 0.051 |
| Less than high school education, geocoded | 18.1 (14.04) | 17.3 (13.34) | 0.65 |
| Tobacco use |  |  | 0.14 |
| Ever | 157 (57.1%) | 286 (51.7%) |  |
| Never | 118 (42.9%) | 267 (48.3%) |  |
| Unknown | 2 (.%) | 1 (.%) |  |
| Diagnosis of alcohol abuse in the past year | 13 (4.7%) | 28 (5.1%) | 0.91 |
| Diagnosis of alcohol abuse any time in the past | 30 (10.8%) | 51 (9.2%) | 0.61 |
| Family history of pancreatic cancer | 18 (6.5%) | 5 (0.9%) | <.001 |
| BMI |  |  | 0.88 |
| Underweight (<18.5) | 7 (2.5%) | 22 (4%) |  |
| Normal weight (18.5-24.9) | 79 (28.5%) | 166 (30%) |  |
| Overweight (25-29.9) | 100 (36.1%) | 187 (33.8%) |  |
| Obese (30+) | 86 (31%) | 169 (30.5%) |  |
| Unknown | 5 (1.8%) | 10 (1.8%) |  |
| Weight change in one year(kg) |  |  | 0.07 |
| Median (IQR) | -2.7 (-5.4, 0.9) | -2.1 (-4.5, 1.5) |  |
| ≤ -6 kg | 50 (18.1%) | 89 (16.1%) |  |
| > -6 & ≤ -4 kg | 23 (8.3%) | 38 (6.9%) |  |
| > -4 & ≤ -2 kg | 34 (12.3%) | 71 (12.8%) |  |
| > -2 & < 2 kg | 87 (31.4%) | 167 (30.1%) |  |
| ≥ 2 & < 4 kg | 22 (7.9%) | 44 (7.9%) |  |
| ≥ 4 kg | 14 (5.1%) | 60 (10.8%) |  |
| Unknown | 47 (17%) | 85 (15.3%) |  |
| Diabetes | 122 (44%) | 191 (34.5%) | 0.004 |
| Biliary tract disease | 58 (20.9%) | 114 (20.6%) | 0.62 |
| Depression | 87 (31.4%) | 181 (32.7%) | 0.78 |
| Deep vein thrombosis | 9 (3.2%) | 37 (6.7%) | 0.08 |
| Gallstone disorders | 47 (17%) | 98 (17.7%) | 0.89 |
| Hereditary cancer syndromes | 37 (13.4%) | 89 (16.1%) | 0.35 |
| Peptic ulcer | 5 (1.8%) | 16 (2.9%) | 0.25 |
| Stage at diagnosis |  |  |  |
| I | 36 (13.0%) |  |  |

|  |  |
| --- | --- |
| II | 60 (21.7%) |
| III | 21 (7.6%) |
| IV | 102 (36.8%) |
| Unknown | 58 (21.0%) |

**Figure 1.**
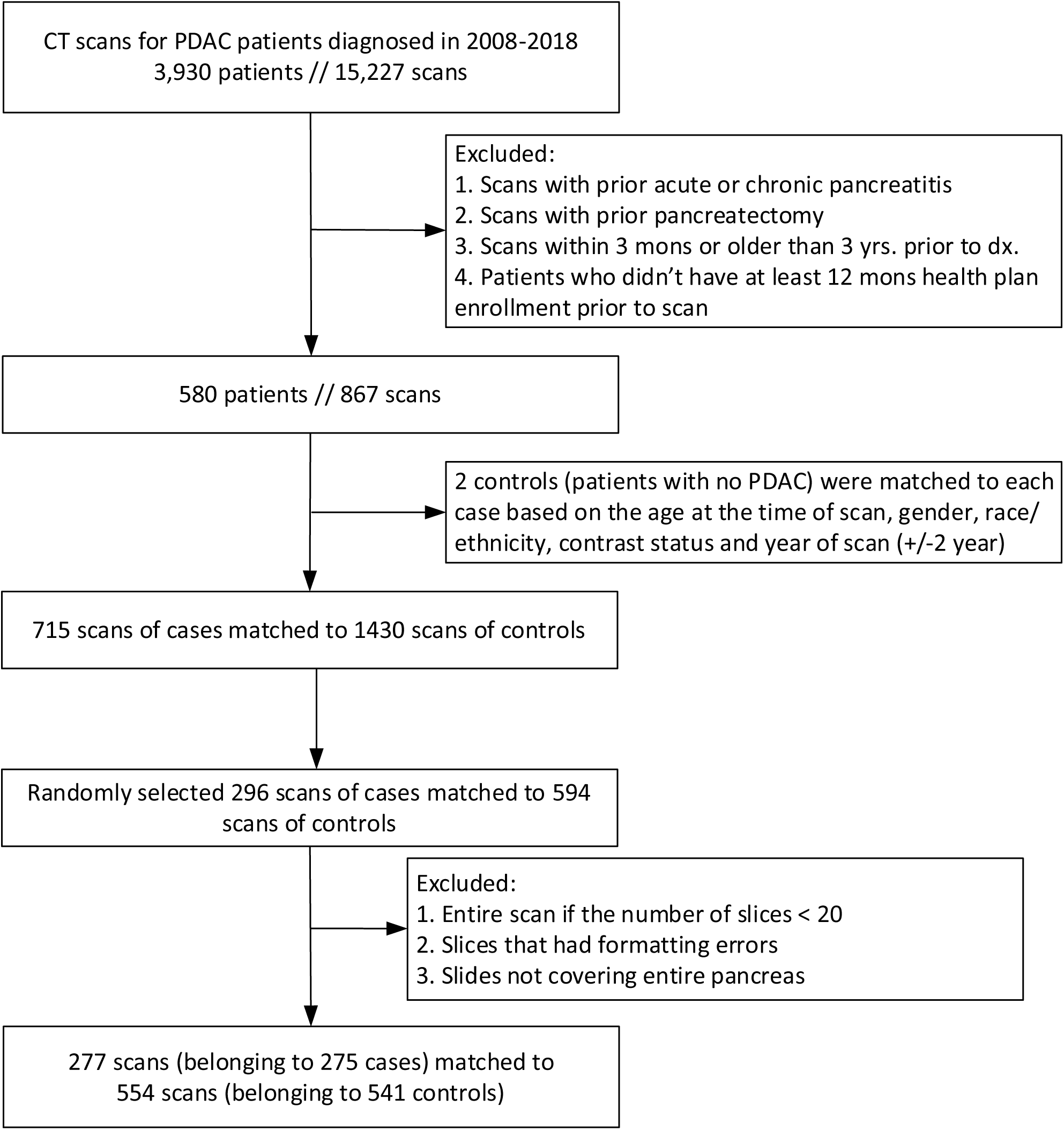
Consort diagram.

### Pancreas Segmentation

The average of DSC from the automated pancreas segmentation algorithm was 83.25 (range ± SD 5.19). Two example images with automated pancreas segmentation and manual group reference segmentation are presented in Figure 2.

**Figure 2.**
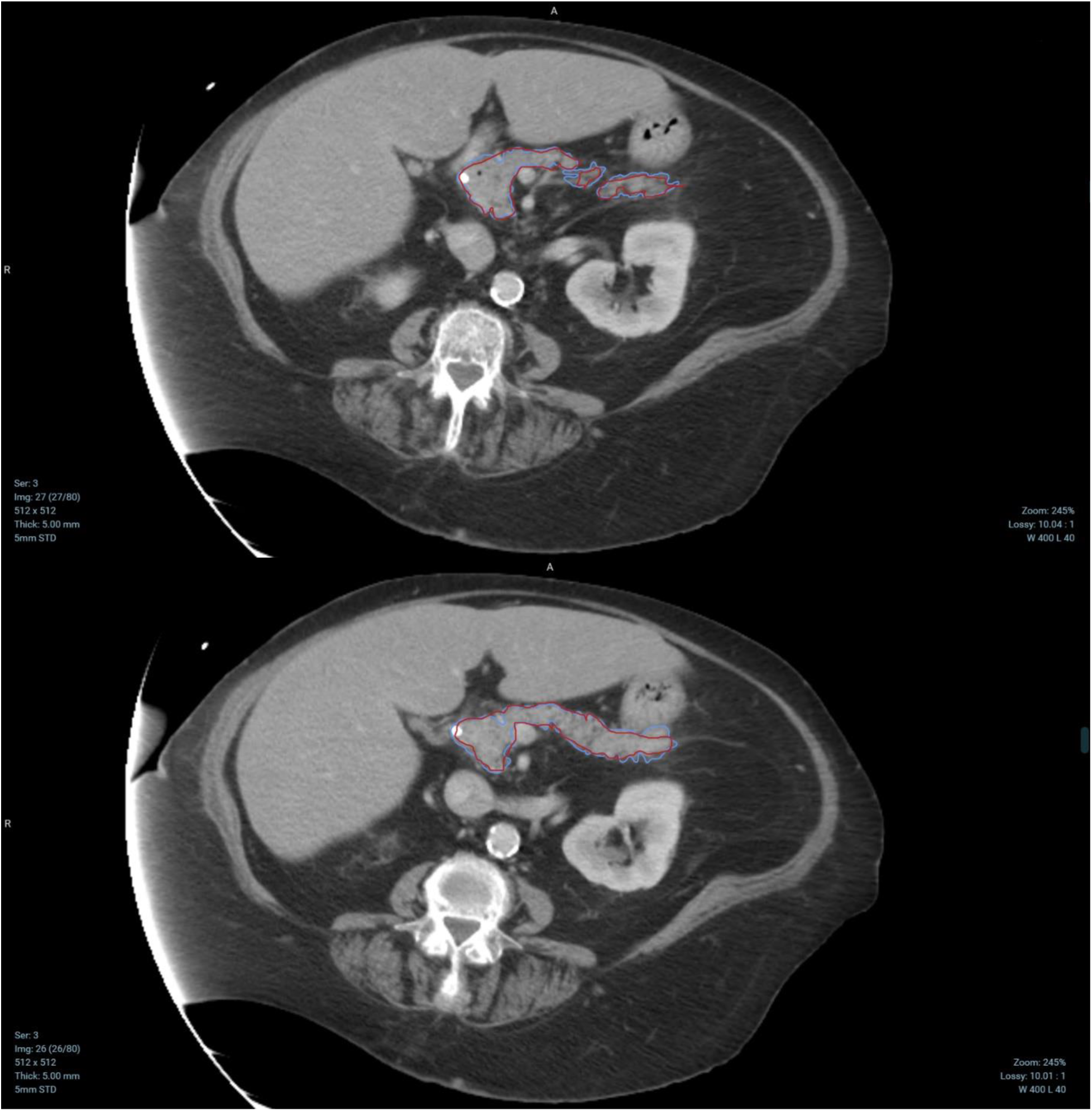
Two example images with automated pancreas segmentation (blue) and manual group truth segmentation (red).

### Model training and validation

For NCA, 5 QIF were selected (Supplemental Document 8). For PCA, 19 principal components were formed (Supplemental Document 9). Tables 2a and 2b show the performance measures of the prediction algorithms based on the QIF selected by NCA and the principal components formed by PCA, respectively.

**Table 2.**
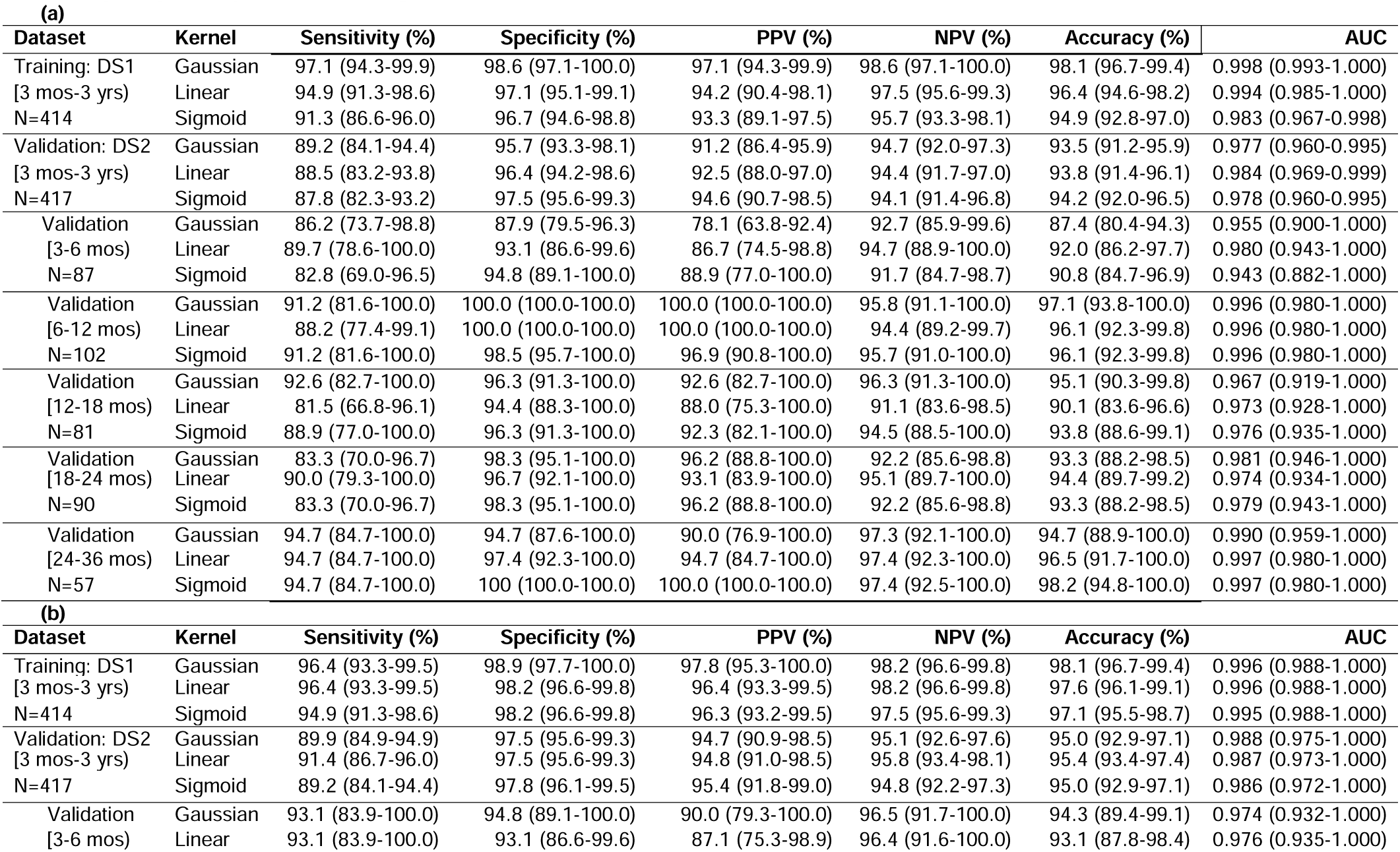
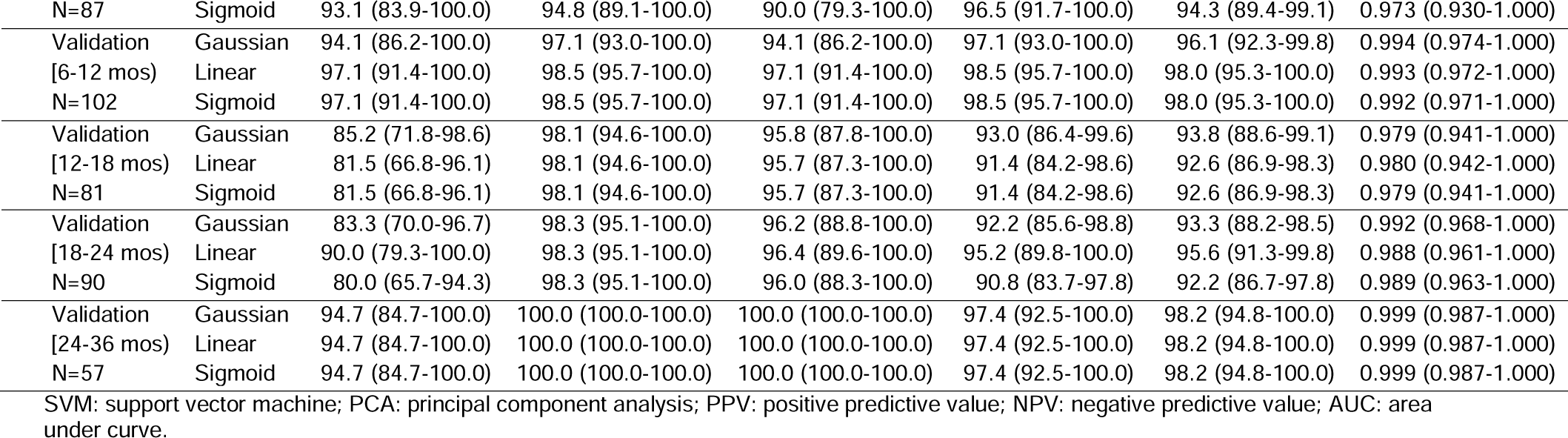
Performance of conditional SVM classifier with various kernel functions based on (a) the 5 features selected by NCA, and (b) the 19 principal components formed by PCA. Percent (%) and 95% confidence interval (CI).

| **(a)** |  |  |  |  |  |  |  |
| --- | --- | --- | --- | --- | --- | --- | --- |
| **Dataset** | **Kernel** | **Sensitivity (%)** | **Specificity (%)** | **PPV (%)** | **NPV (%)** | **Accuracy (%)** | **AUC** |
| Training: DS1 | Gaussian | 97.1 (94.3-99.9) | 98.6 (97.1-100.0) | 97.1 (94.3-99.9) | 98.6 (97.1-100.0) | 98.1 (96.7-99.4) | 0.998 (0.993-1.000) |
| [3 mos-3 yrs] | Linear | 94.9 (91.3-98.6) | 97.1 (95.1-99.1) | 94.2 (90.4-98.1) | 97.5 (95.6-99.3) | 96.4 (94.6-98.2) | 0.994 (0.985-1.000) |
| N=414 | Sigmoid | 91.3 (86.6-96.0) | 96.7 (94.6-98.8) | 93.3 (89.1-97.5) | 95.7 (93.3-98.1) | 94.9 (92.8-97.0) | 0.983 (0.967-0.998) |
| Validation: DS2 | Gaussian | 89.2 (84.1-94.4) | 95.7 (93.3-98.1) | 91.2 (86.4-95.9) | 94.7 (92.0-97.3) | 93.5 (91.2-95.9) | 0.977 (0.960-0.995) |
| [3 mos-3 yrs] | Linear | 88.5 (83.2-93.8) | 96.4 (94.2-98.6) | 92.5 (88.0-97.0) | 94.4 (91.7-97.0) | 93.8 (91.4-96.1) | 0.984 (0.969-0.999) |
| N=417 | Sigmoid | 87.8 (82.3-93.2) | 97.5 (95.6-99.3) | 94.6 (90.7-98.5) | 94.1 (91.4-96.8) | 94.2 (92.0-96.5) | 0.978 (0.960-0.995) |
| Validation | Gaussian | 86.2 (73.7-98.8) | 87.9 (79.5-96.3) | 78.1 (63.8-92.4) | 92.7 (85.9-99.6) | 87.4 (80.4-94.3) | 0.955 (0.900-1.000) |
| [3-6 mos] | Linear | 89.7 (78.6-100.0) | 93.1 (86.6-99.6) | 86.7 (74.5-98.8) | 94.7 (88.9-100.0) | 92.0 (86.2-97.7) | 0.980 (0.943-1.000) |
| N=87 | Sigmoid | 82.8 (69.0-96.5) | 94.8 (89.1-100.0) | 88.9 (77.0-100.0) | 91.7 (84.7-98.7) | 90.8 (84.7-96.9) | 0.943 (0.882-1.000) |
| Validation | Gaussian | 91.2 (81.6-100.0) | 100.0 (100.0-100.0) | 100.0 (100.0-100.0) | 95.8 (91.1-100.0) | 97.1 (93.8-100.0) | 0.996 (0.980-1.000) |
| [6-12 mos] | Linear | 88.2 (77.4-99.1) | 100.0 (100.0-100.0) | 100.0 (100.0-100.0) | 94.4 (89.2-99.7) | 96.1 (92.3-99.8) | 0.996 (0.980-1.000) |
| N=102 | Sigmoid | 91.2 (81.6-100.0) | 98.5 (95.7-100.0) | 96.9 (90.8-100.0) | 95.7 (91.0-100.0) | 96.1 (92.3-99.8) | 0.996 (0.980-1.000) |
| Validation | Gaussian | 92.6 (82.7-100.0) | 96.3 (91.3-100.0) | 92.6 (82.7-100.0) | 96.3 (91.3-100.0) | 95.1 (90.3-99.8) | 0.967 (0.919-1.000) |
| [12-18 mos] | Linear | 81.5 (66.8-96.1) | 94.4 (88.3-100.0) | 88.0 (75.3-100.0) | 91.1 (83.6-98.5) | 90.1 (83.6-96.6) | 0.973 (0.928-1.000) |
| N=81 | Sigmoid | 88.9 (77.0-100.0) | 96.3 (91.3-100.0) | 92.3 (82.1-100.0) | 94.5 (88.5-100.0) | 93.8 (88.6-99.1) | 0.976 (0.935-1.000) |
| Validation | Gaussian | 83.3 (70.0-96.7) | 98.3 (95.1-100.0) | 96.2 (88.8-100.0) | 92.2 (85.6-98.8) | 93.3 (88.2-98.5) | 0.981 (0.946-1.000) |
| [18-24 mos] | Linear | 90.0 (79.3-100.0) | 96.7 (92.1-100.0) | 93.1 (83.9-100.0) | 95.1 (89.7-100.0) | 94.4 (89.7-99.2) | 0.974 (0.934-1.000) |
| N=90 | Sigmoid | 83.3 (70.0-96.7) | 98.3 (95.1-100.0) | 96.2 (88.8-100.0) | 92.2 (85.6-98.8) | 93.3 (88.2-98.5) | 0.979 (0.943-1.000) |
| Validation | Gaussian | 94.7 (84.7-100.0) | 94.7 (87.6-100.0) | 90.0 (76.9-100.0) | 97.3 (92.1-100.0) | 94.7 (88.9-100.0) | 0.990 (0.959-1.000) |
| [24-36 mos] | Linear | 94.7 (84.7-100.0) | 97.4 (92.3-100.0) | 94.7 (84.7-100.0) | 97.4 (92.3-100.0) | 96.5 (91.7-100.0) | 0.997 (0.980-1.000) |
| N=57 | Sigmoid | 94.7 (84.7-100.0) | 100 (100.0-100.0) | 100.0 (100.0-100.0) | 97.4 (92.5-100.0) | 98.2 (94.8-100.0) | 0.997 (0.980-1.000) |
| **(b)** |  |  |  |  |  |  |  |
| **Dataset** | **Kernel** | **Sensitivity (%)** | **Specificity (%)** | **PPV (%)** | **NPV (%)** | **Accuracy (%)** | **AUC** |
| Training: DS1 | Gaussian | 96.4 (93.3-99.5) | 98.9 (97.7-100.0) | 97.8 (95.3-100.0) | 98.2 (96.6-99.8) | 98.1 (96.7-99.4) | 0.996 (0.988-1.000) |
| [3 mos-3 yrs] | Linear | 96.4 (93.3-99.5) | 98.2 (96.6-99.8) | 96.4 (93.3-99.5) | 98.2 (96.6-99.8) | 97.6 (96.1-99.1) | 0.996 (0.988-1.000) |
| N=414 | Sigmoid | 94.9 (91.3-98.6) | 98.2 (96.6-99.8) | 96.3 (93.2-99.5) | 97.5 (95.6-99.3) | 97.1 (95.5-98.7) | 0.995 (0.988-1.000) |
| Validation: DS2 | Gaussian | 89.9 (84.9-94.9) | 97.5 (95.6-99.3) | 94.7 (90.9-98.5) | 95.1 (92.6-97.6) | 95.0 (92.9-97.1) | 0.988 (0.975-1.000) |
| [3 mos-3 yrs] | Linear | 91.4 (86.7-96.0) | 97.5 (95.6-99.3) | 94.8 (91.0-98.5) | 95.8 (93.4-98.1) | 95.4 (93.4-97.4) | 0.987 (0.973-1.000) |
| N=417 | Sigmoid | 89.2 (84.1-94.4) | 97.8 (96.1-99.5) | 95.4 (91.8-99.0) | 94.8 (92.2-97.3) | 95.0 (92.9-97.1) | 0.986 (0.972-1.000) |
| Validation | Gaussian | 93.1 (83.9-100.0) | 94.8 (89.1-100.0) | 90.0 (79.3-100.0) | 96.5 (91.7-100.0) | 94.3 (89.4-99.1) | 0.974 (0.932-1.000) |
| [3-6 mos] | Linear | 93.1 (83.9-100.0) | 93.1 (86.6-99.6) | 87.1 (75.3-98.9) | 96.4 (91.6-100.0) | 93.1 (87.8-98.4) | 0.976 (0.935-1.000) |
| N=87 | Sigmoid | 93.1 (83.9-100.0) | 94.8 (89.1-100.0) | 90.0 (79.3-100.0) | 96.5 (91.7-100.0) | 94.3 (89.4-99.1) | 0.973 (0.930-1.000) |
| Validation | Gaussian | 94.1 (86.2-100.0) | 97.1 (93.0-100.0) | 94.1 (86.2-100.0) | 97.1 (93.0-100.0) | 96.1 (92.3-99.8) | 0.994 (0.974-1.000) |
| [6-12 mos) | Linear | 97.1 (91.4-100.0) | 98.5 (95.7-100.0) | 97.1 (91.4-100.0) | 98.5 (95.7-100.0) | 98.0 (95.3-100.0) | 0.993 (0.972-1.000) |
| N=102 | Sigmoid | 97.1 (91.4-100.0) | 98.5 (95.7-100.0) | 97.1 (91.4-100.0) | 98.5 (95.7-100.0) | 98.0 (95.3-100.0) | 0.992 (0.971-1.000) |
| Validation | Gaussian | 85.2 (71.8-98.6) | 98.1 (94.6-100.0) | 95.8 (87.8-100.0) | 93.0 (86.4-99.6) | 93.8 (88.6-99.1) | 0.979 (0.941-1.000) |
| [12-18 mos) | Linear | 81.5 (66.8-96.1) | 98.1 (94.6-100.0) | 95.7 (87.3-100.0) | 91.4 (84.2-98.6) | 92.6 (86.9-98.3) | 0.980 (0.942-1.000) |
| N=81 | Sigmoid | 81.5 (66.8-96.1) | 98.1 (94.6-100.0) | 95.7 (87.3-100.0) | 91.4 (84.2-98.6) | 92.6 (86.9-98.3) | 0.979 (0.941-1.000) |
| Validation | Gaussian | 83.3 (70.0-96.7) | 98.3 (95.1-100.0) | 96.2 (88.8-100.0) | 92.2 (85.6-98.8) | 93.3 (88.2-98.5) | 0.992 (0.968-1.000) |
| [18-24 mos) | Linear | 90.0 (79.3-100.0) | 98.3 (95.1-100.0) | 96.4 (89.6-100.0) | 95.2 (89.8-100.0) | 95.6 (91.3-99.8) | 0.988 (0.961-1.000) |
| N=90 | Sigmoid | 80.0 (65.7-94.3) | 98.3 (95.1-100.0) | 96.0 (88.3-100.0) | 90.8 (83.7-97.8) | 92.2 (86.7-97.8) | 0.989 (0.963-1.000) |
| Validation | Gaussian | 94.7 (84.7-100.0) | 100.0 (100.0-100.0) | 100.0 (100.0-100.0) | 97.4 (92.5-100.0) | 98.2 (94.8-100.0) | 0.999 (0.987-1.000) |
| [24-36 mos) | Linear | 94.7 (84.7-100.0) | 100.0 (100.0-100.0) | 100.0 (100.0-100.0) | 97.4 (92.5-100.0) | 98.2 (94.8-100.0) | 0.999 (0.987-1.000) |
| N=57 | Sigmoid | 94.7 (84.7-100.0) | 100.0 (100.0-100.0) | 100.0 (100.0-100.0) | 97.4 (92.5-100.0) | 98.2 (94.8-100.0) | 0.999 (0.987-1.000) |

SVM: support vector machine; PCA: principal component analysis; PPV: positive predictive value; NPV: negative predictive value; AUC: area under curve.

#### Performance based on entire scan window (3 mos-3 yrs prior to t_0_)

Accuracy/AUC based on DS2 were 94%/0.98 and 95%/0.99, respectively, for prediction algorithms developed based on NCA and PCA, regardless of kernel functions applied. Sensitivity, specificity, PPV, and NPV based on DS2 were in the ranges of 88-91%, 96-98%, 91-95%, and 94-96%, varying slightly by QIF selection method (NCA vs. PCA) and kernel function used to develop the prediction algorithms. Algorithms developed based on PCA had better performance than those of NCA for all measures regardless of kernel functions applied.

#### Performance based on 5 individual sub-time windows

The performance within each of the time windows remained high for both NCA and PCA methods. For the PCA method, the lowest sensitivity, specificity, PPV, NPV, accuracy, and AUC were 82%, 93%, 87%, 91%, 93%, and 0.97, respectively. QIF within 2-3 years prior to t_0_ also had very high predictive power (accuracy 95-98%; AUC 0.99-1.00).

### Exploratory Analyses

6 and 14 QIF were selected from EDS1 and EDS2, respectively (Supplemental Document 6, Tables S1 and S3). Five of the 6 QIF selected from EDS1 were in the list of QIF selected from EDS2. The performance measures between the two sets of selected QIF were similar (Supplemental Document 6, Tables S2 and S4). The performance measures between the two sets of data were comparable.

### Clinical evaluation

In the 19 PDAC cases with pre-diagnostic images in 24-36 months, 14, 4 and 1 patients were classified as having low, medium, and high risk based on the manual review of the CT images, respectively (Table 3). Similarly, in the 19 matched controls, an overwhelming majority were believed to have low risk based on the manual review of the CT images (Table 3). However, the computer algorithms only mistakenly predicted one patient in cases and correctly classified all the controls.

**Table 3.** A blind^a^ manual review of pre-diagnostic CT images in 24-36 months prior to PDAC diagnosis and the CT images of the matched controls by case-control status and compute labels (high risk vs. low risk)^b^.

| Manually assigned<sup>c</sup> risk of PDAC | Cases (n=19) |  |  | Controls (n=19) |  |  |
| --- | --- | --- | --- | --- | --- | --- |
|  | Labeled by computer as having high risk (n=18) | Labeled by computer as having low risk (n=1) | Total | Labeled by computer as having high risk (n=0) | Labeled by computer as having low risk (n=19) | Total |
| Low | 14 | 0 | 14 | 0 | 16 | 16 |
| Median | 3 | 1 | 4 | 0 | 2 | 2 |
| High | 1 | 0 | 1 | 0 | 0 | 0 |
| Non-diagnostic | 0 | 0 | 0 | 0 | 1 | 1 |

a: Reviewer was not informed about the computer labels or PDAC case/control status at the time of review.
b: Computer label was assigned according to the prediction algorithm developed with Gaussian kernel function and principal component analysis.

## DISCUSSION

In this study, we developed and validated automated computer algorithms to predict PDAC solely based on QIF of pre-diagnostic CT scans. Our results showed QIF of pre-diagnostic scans can accurately predict PDAC in 3-36 months prior to diagnosis. Interestingly, when the validation was stratified by timing of scan in relation to diagnosis, the performance seemed to be maintained in all the time periods examined prior to cancer diagnosis. The QIF-based algorithm had excellent ability to predict PDAC (sensitivity 95%, specificity 100%, PPV 100%, NPV 97%, accuracy 98% and AUC 100%) based on scans from 24-36 months prior to cancer diagnosis. A manual assessment of the scans in 24-36 months prior to PDAC diagnosis revealed that an overwhelming majority had low risk of PDAC, yet the computer algorithm was able to predict the outcome correctly except for one case who had mild ductal prominence and loss of lobulation in pancreas head.

A major barrier for early detection in pancreatic cancer is the inability to reliably identify precursor lesions based on conventional imaging. Pancreatic Intra-epithelial neoplasia (PanIN) III or high-grade dysplasia are histologic findings not typically visible on cross-sectional imaging^36^. Previous studies have identified specific abnormalities that can be identified in up to 50% of cancer cases prior to diagnosis of PDAC including main duct dilatation, atrophy and duct stricture.^4,37,38^ However, such findings lack sensitivity and in many cases are subject to interpreter variability. Therefore, a systematic approach of direct imaging analysis that applies objective assessment of imaging-based parameters provides a promising opportunity to identify imaging-based signatures of early pancreatic cancer. The performance of the algorithms was high in all the time windows being studied.

Radiomics has been applied in the diagnosis, prediction, and prognosis of other cancer types (e.g. lung cancer, lung nodule and breast cancer).^39^ While radiomics has been applied for prognosis in PDAC^7-12^, very little progress has been made with respect to early detection. Using 225 training and 125 validation images, Chu et al. differentiated CT scans of PDAC patients and healthy controls with a sensitivity of 100% and specificity 98.5%.^18^ However, in Chu’s study, the images being analyzed were taken after diagnosis (mean tumor size 4.1 cm). In addition, the pancreas boundaries were manually segmented^18^ and thus, utility of the algorithm in clinical operation is limited. In the current study, pre-diagnostic scans were applied, and the segmentation of pancreas was computerized.

A key distinction related to assessment of pre-diagnostic imaging is whether the automated QIF algorithms were classifying cancer risk based on features readily visible on the images or incorporating additional elements from the data contained within the images. What we observed in the current study is that relying on human assessment of the pre-diagnostic CT images taken 2-3 years prior to cancer diagnosis by an expert body-image radiologist with 15 years of experience failed to distinguish cancer and healthy patients at the level of performance obtained by the radiomics-based algorithm suggesting the QIF provide additional information beyond identification of established radiographic findings.

The current study has several strengths. First, we implemented a pancreas segmentation algorithm and series of quantitative imaging features that were previously validated. Therefore, the developed classifiers can potentially be widely implemented across health care systems. Second, we relied on pre-diagnostic images 3 months-3 years prior to cancer diagnosis. Thus, the developed algorithms have the potential to predict pancreatic cancer in a time frame that would allow for sufficient lead-time for intervention to impact the disease course. Third, we applied two machine learning approaches to select the most influential radiomic features. Although both methods worked well, it appears that algorithms based on principal components formed by PCA are more accurate. This is likely to be the result of the more inclusive nature of PCA compared to NCA. Finally, the comparison between blinded expert human review and automated QIF-based algorithm helped to demonstrate the potential added-value of this approach beyond identification of established abnormalities of the pancreatic parenchymal or duct system.

The current study had several limitations. First, although the images being used in the analyses were at least 3 months prior to cancer diagnosis, a small number of scans may contain visible tumors, which may deform the shape of pancreas and thus negatively impact the performance of the final algorithms. A few (< 5) images were manually removed due to visible tumors.

However, additional images with visible tumors may still have been present. Second, when the weights of the convolutional neural network for pancreas segmentation derived previously were adjusted to fit the scans of the current study, the process was only applied to the scans of cases but not to the scans of controls. Third, we did not consider higher-order statistics QIF (e.g. Gabor wavelet transformation) in the study. Previously studies have shown that mapping QIF into a higher-order feature space can further improve accuracy.^13,15^ Fourth, cases and controls were not matched by the indications for the scans. Fifth, the manual assessment for PDAC risk was only performed for images in 2-3 years prior to the index date. Finally, the current study lacks an external validation, in which CT images from another health care may be used to test the robustness of our algorithms.

## CONCLUSION

The radiomics-based automated algorithms provide a method to detect PDAC as early as 2-3 years prior to cancer diagnosis. The algorithm has the potential to be used for future early detection protocols for pancreatic cancer. Future studies are needed to understand feasibility, challenges, and cost-effectiveness of such an implementation.

## Data Availability

Anonymized data that support the findings of this study may be made available from the investigative team in the following conditions: 1) agreement to collaborate with the study team on all publications, 2) provision of external funding for administrative and investigator time necessary for this collaboration, 3) demonstration that the external investigative team is qualified and has documented evidence of training for human subjects protections, and 4) agreement to abide by the terms outlined in data use agreements between institutions.

## Abbreviations

AUC: area under the curve
CI: confidence interval
CT: computerized tomography
DSC: Dice similarity coefficient
NCA: neighborhood component analysis
NOD: new onset diabetes
NPV: negative predictive value
KPSC: Kaiser Permanente Southern California
SEER: Surveillance, Epidemiology, and End Results
ICD-10-CM: Tenth Revision of International Classification of Diseases, Clinical Modification
PCA: principal component analysis
PDAC: pancreatic ductal adenocarcinoma
PPV: positive predictive value
QIF: quantitative imaging features
SVM: support vector machine

## Acknowledgement

The authors thank Sole Cardoso for the assistance with formatting the manuscript, and Botao Zhou for the additional analyses. The images were kindly provided by the Medical Imaging Technology and Informatics group of Kaiser Permanente Southern California Permanente Medical Group.

